# Hospital bed capacity across in Tunisia hospital during the first four waves of the COVID-19 pandemic: A descriptive analysis

**DOI:** 10.1101/2022.08.23.22279122

**Authors:** Slimane BenMiled, Chiraz Borgi, Mohamed Hsairi, Naoufel Somrani, Amira Kebir

## Abstract

**Background:** On March 11, 2020, WHO announced that the COVID-19 epidemic had passed the pandemic stage, indicating its spread over several continents. Tunisia’s containment and targeted screening strategy corresponded to the first WHO guidelines. Since then, public health policy has been more flexible and focused on the management of hospital beds.

**Objective:** Our aims are to analyze bed occupancies for public hospitals and time delay reponse from the health care system in regard to the epidemiological situation.

**Methods:** We have analyzed the evolution of daily cases in relation to the different NPI actions undertaken by the Tunisian Government between March 2019 and February 2022 using the CoMo model. We have also studied the flexibility of the O2 and ICU public hospital bed occupancies. We have used three distinct indices to assess this flexibility: the Ramp Duration Until the Peak (RDUP), which measures the duration of the wave/bed allocation effort. The Ramp Growth Until the Peak (RGUP), measures the peak height, and Ramp Rate Until the Peak (RRUP) measures the growth rate of the wave. Also, in order to evaluate the government response efficacy, we have calculated the time delay at the start (resp. at peak) of each two waves.

**Results:**

1. The evolution of the epidemic in Tunisia was divided into two phases, the first of which corresponded to the initial wave, during which the pandemic was controlled due to very strong NPI actions. The second phase was distinguished by a progressive relaxation of measures and an increase in wave intensity.
2. ICU bed availability has followed the demand for beds, while ICU bed occupancy has always been higher than 85% with a maximum of 97%.
3. In terms of bed distribution, the government’s response was slow (9.4 days for the 02 beds and 18.2 days for the ICU beds). The same may be said for the reaction in terms of bed reallocation in the original departments (16 days for ICU beds and 10.6 days for O2 beds)..

**Conclusion:** We were able to examine the responsiveness of the system as a whole for all of Tunisia’s public hospitals by measuring the flexibility and bed margin. With this research, decision makers will be able to assess their response capabilities in the event of current pandemic, as well as a future one.

## 1 Introduction

On 30 January 2020, COVID-19 was retained by the World health organization (WHO) as a public health emergency of international concern; and WHO announced the pandemic threat on March 11, 2020, ((Sohrabi et al., 2020; WHO, 2020)).

In Tunisia, the first COVID-19 wave appear on March 2, 2020, followed by several implemented preventive measures to reduce transmission levels among the population. These measures included mask use, physical distancing, school and university closures, sports and cultural events ban, borders closure, targeted screening, and finally a national lockdown announced on March 22, 2020, ((Ouchetto et al., 2020)). However, in July 2020, Non-pharmaceutical interventions (NPI) measures have been significantly reduced. This resulted in more intense waves beginning in October 2020, forcing the Tunisian Ministry of Health to respond rapidly by strengthening its resources, particularly in terms of hospital management.

In these conditions, better healthcare administration becomes a crucial concern Didier ((2016)). Flexibility in hospital bed management, in particular, has become critical in treating patients with severe and/or serious COVID-19 infections ((Bekker et al., 2017)).

The goal of this study is to describe the COVID-19 situation between July 2020 and February 2022, by using the data from the Facebook coronavirus survey ^1^, Health Metric Data^2^ and using newspaper investigation. We also investigate hospital bed occupancies (beds for patients who require oxygen / ICU beds) at the national level. On the other hand, a mathematical index was developed and used to analyze the time delay between daily cases and mortality, as well as bed occupancy.

In section 2, we go over the databases used in this study, as well as the methodology and tools used to analyze the data. We present our main results in section 3 which we discuss in section 4. Finally, in section 5 a conclusion is presented.

## 2 Materials and methods

### 2.1 Data collection method

#### 2.1.1 Bed occupancy data

Bed occupancy was obtained for public Tunisian hospitals based on data from the COVID-19 SHOC Room (Strategic Health Operations Center Room) (SR) and the Ministry of Health’s Computer Centre (CIMS). The goal of the SR was to collect daily data from hospitals, communicate with them to filter and debug mistakes, and illustrate the results. The data was collected daily using a spreadsheet from 168 hospitals located throughout Tunisia’s 24 regions. The CIMS is in charge of running the administrative management system for admissions and exits in hospitals with an IT management system. Specifically, the 168 hospitals in the SR database.

Collected data ranked hospital beds (*i*.*e*. beds that had all the equipment and personnel necessary for their function) according to two large groups, ICU beds, and non-ICU beds called here O2 beds; ICU beds are used in intensive care units (ICUs) to treat patients with serious or life-threatening illnesses and traumas. ICUs differ from ordinary hospital wards in that they have a higher staff-to-patient ratio and have access to advanced medical resources and equipment not commonly found elsewhere. The beds assigned to each group were further subdivided into available beds, occupied beds (beds that were occupied by inpatients), and unoccupied beds (*i*.*e*., available beds that were not occupied).

For our analysis, we collected the following daily reported data stream: (i) The SR database contains data on O2 dedicated beds, O2 occupied beds, O2 available beds, ICU dedicated beds, ICU occupied beds and ICU available beds. (ii) The CIMS database that contains data on admissions and exits date for each COVID-19 patient, the hospital unit of entrance and exit, and its status at existing (*i*.*e*. death or not). Both SR and CIMS databases cover the period from September 2020 to the end of February 2022. The data have been cured and aggregated at the national level. Database is included in the appendix B and available at github : https://github.com/slimane66/bedOccupancyTunisia.git.

During this period the mean occupied 02 bed was 972 and the mean occupied ICU bed was 240 (see figures 1 and table 1).

**Figure 1:**
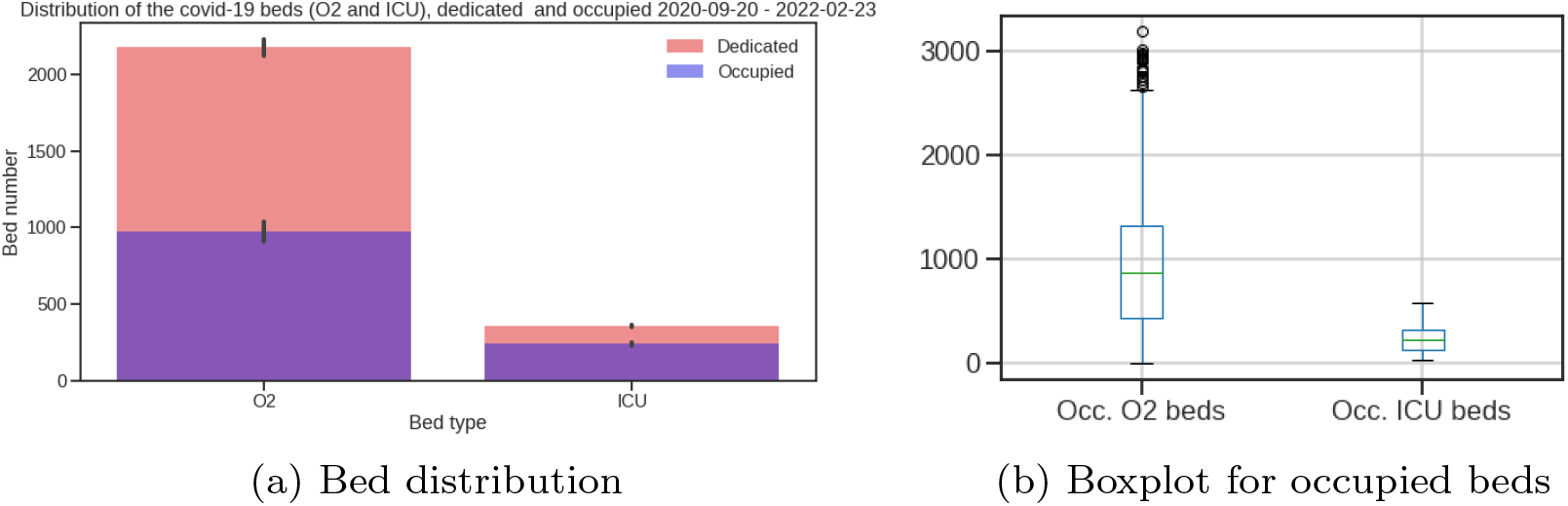
Distribution of hospital beds by bed type: O2/ICU beds dedicated/occupied

**Table 1:** Average, min and max Bed occupancy

|  | O2 |  |  | ICU |  |  |
| --- | --- | --- | --- | --- | --- | --- |
|  | average | min | max | average | min | max |
| Occupancy | 42% | 2% | 90% | 66% | 10% | 97% |
| Date |  | 18/11/2021 | 07/07/2021 |  | 11/11/2021 | 26/07/2021 |
| Dedicated | 2175 | 2410 | 2602 | 359 | 361 | 544 |
| Occupied | 972 | 61 | 2367 | 240 | 37 | 528 |

In the remainder of the document, we will use the term *bed curves* or *bad waves* for curves and waves relating to data on allocated or occupied beds ICU or O2.

Since both the epidemiological and bed occupancy data are noisy, we smoothed them using a linear convolution one dimension kernel, *Box*1*DKernel*(*N*) with *N* = 7 days from *astropy*.*convolution*((Robitaille et al., 2013)) library on Python 3 ((Van Rossum and Drake, 2009)).

### 2.2 Data analysis methodology

#### 2.2.1 Bed occupancy and flexibility

According to Green and V. ((2002)), the ideas of bed occupancy and flexibility were utilized to study the reaction of bed demand. Bed occupancy is defined as the ratio of occupied beds to the total number of allocated COVID-19 beds.

To analyze bed occupancy during the epidemy, we suggest three distinct indices to assess its flexibility: the Ramp Duration Until the Peak (RDUP), which measures the duration of the wave/bed allocation effort. The Ramp Growth Until the Peak (RGUP), which measures the peak height, and Ramp Rate Until the Peak (RRUP) which measures the growth rate of the wave (*i*.*e*. 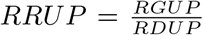) (see figure 8 in supplementary material). In the case of bed allocation curves, RRUP measure the intensity of the bed allocation effort.

The RDUP can be easily computed from the difference between the time where the peak is reached *t*_*p*_ and the starting time of the increase of the wave (*i*.*e*. the start of the wave), *t*_0_. RDUP corresponds to the time to reach the curve peak. In the same way, the RGUP can be obtained from the difference between the number of available beds at the peak *n*_*p*_ and at the starting time of the increase in the demand *n*_0_.

To match different waves, we also calculate the time delay at the start (resp. at peak) of each two wave (see figure 9 in supplementary material).

##### Estimation the wave start and the peak

The classical definition of the start of an epidemiological wave involves the evaluation of the effective reproduction number, *R*_*t*_, defined as the average number of secondary cases per infectious case in a population made up of both susceptible and non-susceptible hosts ((Lash et al., 2020)).

Regardless of the classical definition of the effective reproduction number, *R*_*t*_, its utility stems from the fact that it reveals the curve’s exponential growth. To be more specific, if *R*_*t*_ *>* 1, the curve grows exponentially, whereas if *R*_*t*_ *<* 1, the curve shrinks to zero. As a result, the wave’s beginning and peak correspond to the time when *R*_*t*_ = 1. ((Diekmann et al., 1990)).

Thus, we estimate the start of a wave as the time, *t*_0_, when 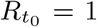 while increasing in a neighborhood of *t*_0_. Similarly the peak corresponds to the time, *t*_*p*_, for which 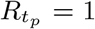 while decreasing in a neighborhood of *t*_*p*_.

When applied to bed occupancy data, similarly to an epidemiological curve, *R*_*t*_ evaluates the exponential growth of the bed occupancy data curve. To avoid any misunderstanding, we’ll refer to it as a ‘gradient like index,’ or *G*_*t*_. Therefore, we define the wave period using *R*_*t*_ or *G*_*t*_ depending on the data type, daily cases or bed occupancy. We computed *R*_*t*_ and *G*_*t*_ using ((Cori et al., 2013)).

### 2.3 Mathematical modelling

#### 2.3.1 Model description

We adapted the CoMo model ((Aguas et al., 2020)) to simulate the spread of SARS-CoV2 in the context of NPIs measures and vaccination strategies. The CoMo model is a dynamic SEIRS (Susceptible-Exposed-Infected-Recovered-Susceptible) model. It is an age-structured SEIRS model with infected compartments stratified by symptoms, severity, treatment-seeking, and hospital access (see figure 10).

The description of the variables used and a list with all parameters included in the full model are given in the supplementary material.

#### 2.3.2 Model calibration

The CoMo model was adapted to the Tunisian context using daily cases and mortality data, demographic data from the National Agency for Statistics of Tunisia^3^, information on the different types of NPI intervention measures carried out (*e*.*g*. school closer, social distance, international travel ban, mask-wearing, …) and parameters on hospitalizations and vaccination in Tunisia (see supplementary material).

Non-pharmaceutical interventions (NPI) measures were collected from Facebook coronavirus survey, Health Metric Data, and using newspaper investigation.

For further exploration of NPI effectiveness, we compared the timeline of NPI implementation with an external data source, the Oxford COVID-19 Government Response Stringency index Hale et al. ((2020)), a quantitative measure of the strictness of government policies regulating population behavior.

Daily cases and deaths were collected from the Official COVID-19 report from WHO database^4^.

The model was calibrated using the epidemic data in Tunisia between September 15, 2020, and February 15, 2022, and tested through June 2022. Parameters that could not be evaluated were estimated by the optimal model fit to epidemiological data.

## 3 Results

### 3.1 Reconstruction of the epidemy history in Tunisia

From March 2020 to February 2022, Tunisia had six pandemic waves. The first one was held from March to June 2020, the second from September to October 2020, and the third from December to January 2021. The historical variant was the most common virus during the first three waves. The alpha variant caused the fourth wave, which occurred between March and May. The delta variant generated the fifth wave, which occurred between July and August. The Omicron variant produced the last wave, which lasted from December 2021 to January 2022 (see figure 2 and supplementary database) ^5^. These six waves were becoming increasingly large.

**Figure 2:**
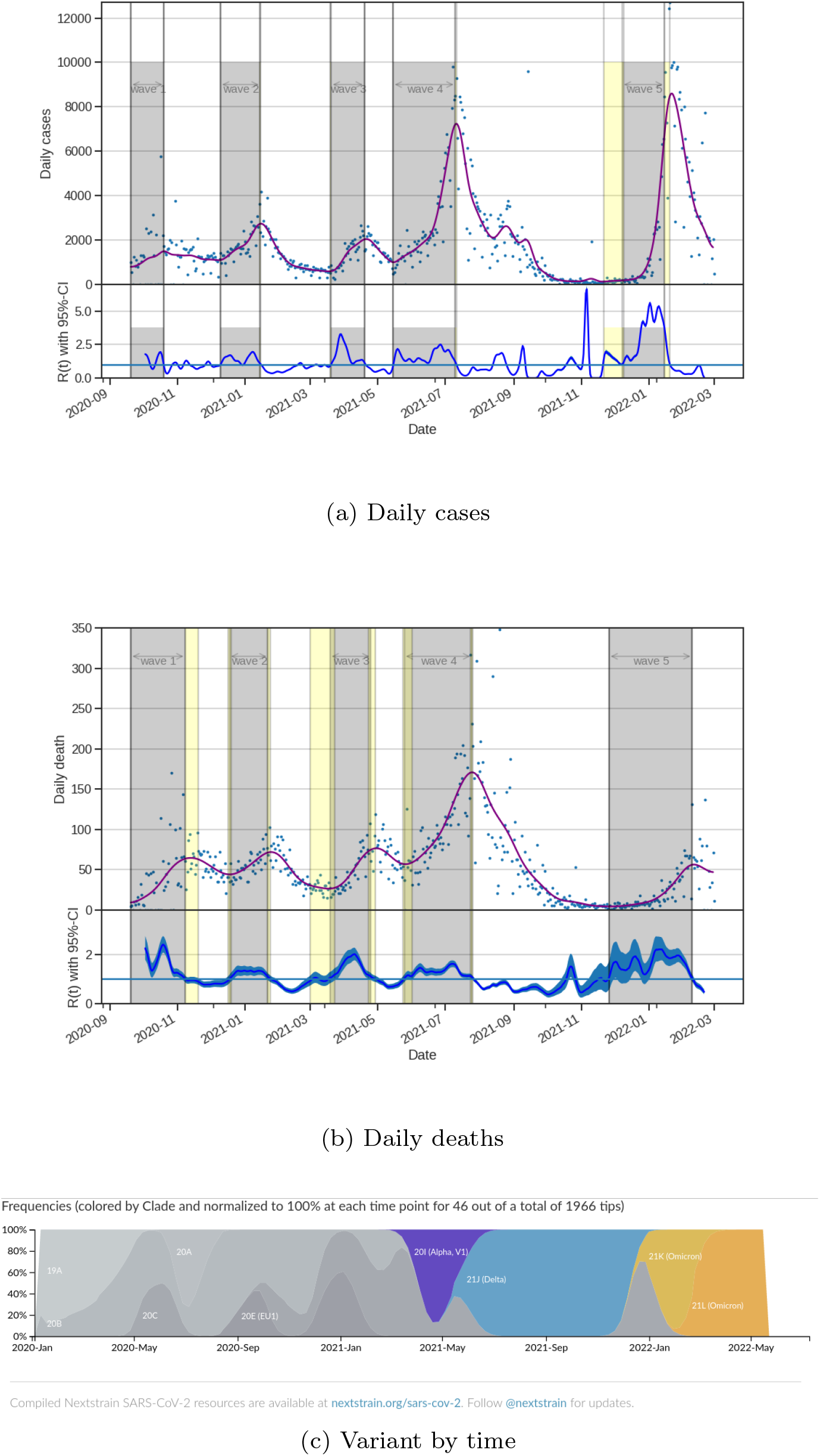
Daily cases, daily deaths with their corresponding *R*_*t*_ and variant proportion from Mars 2019 to February 2022. The grey areas correspond to the different waves. We observe, that the waves have been more and more intens. Variant figure taken from nextstrain.org/sars-cov-2((Hadfield et al., 2018))

By march, 28th, 2022, a total of 1, 033, 731 confirmed cases and 28, 165 deaths due to COVID-19 were recorded.

The Oxford Stringency Index showed the first phase of growth from March to June 2020, followed by a second phase from October 2020 (see figures 3). Since then, the Oxford Stringency Index has been rather stable, ranging between 60 and 100 % during school holidays. Furthermore, except for the first and second waves, no variation in the stringency index occurs before the epidemic’s peak. For example, during the second wave, a 12-hour curfew was imposed at the peak. The decline of this second wave appears to have been caused by regional NPI efforts between September and October 2020, as well as an increase in mask-wearing (see figure 3).

**Figure 3:**
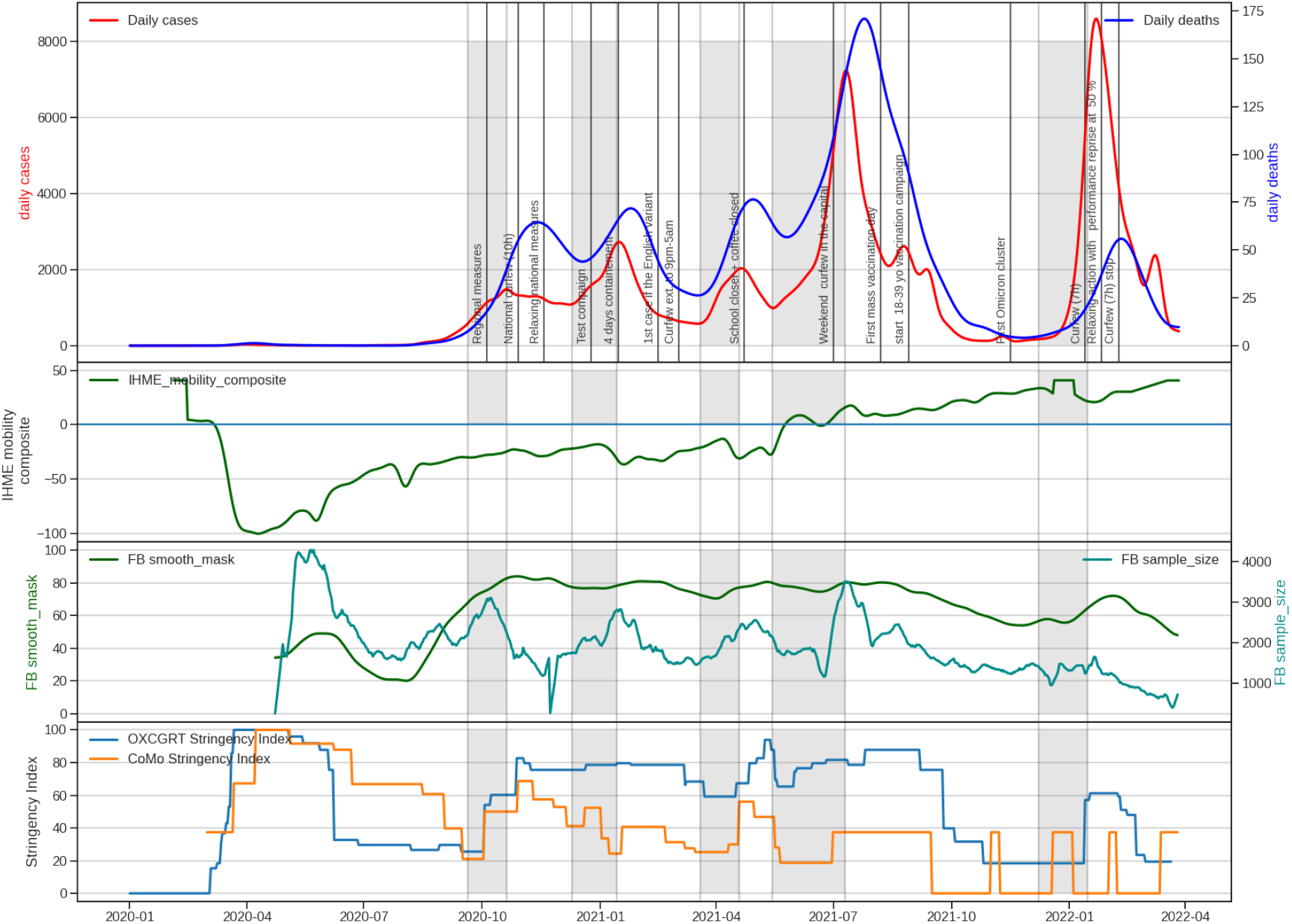
Evolution of COVID19 in Tunisia. From the top to buttom: (1) Daily confirmed case incidence across Tunisia. (2) Facebook coronavirus mask wearing survey. (3) The mobility index. (4) The Oxford NPI stringency index and stringency index calculate with NPI values used in the CoMo model. We observe, that the NPI measures have been less and less followed despite strong NPI decisions.

To understand the response to the NPI actions, we plotted IHME mobility composite between October 2020 and February 2022 (see figure 3). We observe that after a decrease in the mobility index between March and September 2020, the mobility index began to increase and turn to be positive from May 2021. From that date on, people’s mobility was higher than in previous years. Therefore, NPI decisions undertaken by the government to limit movement had a limited effect, and even from May 2021, these decisions had almost no effect.

During the second wave, between August and October 2020, the mask-wearing curve shows a significant increase. It was followed by a period of little change in the values (*<* 20%) until September 2021. During the start of the wave, we observe an increase in both mask use and the number of responses to the inquiry. It’s worth noting that the number of participants in the questionnaire has decreased significantly since the fourth wave, which could skew the results starting in July 2021.

To better understand the effect of the actions taken, we simulated the NPI actions using the CoMo model. We then evaluated the outcome of the NPI measures by minimizing the model output with the observed epidemiological data (*e*.*g*. daily cases and daily deaths). The simulation of the daily cases and daily deaths showed that the model accurately depicts the observed data (see figure 10).

We then calculated the Oxford Stringency Index on the NPI action used to calibrate the model (figure 3).

### 3.2 Bed occupancy analysis

From March 2020 to February 2022, the average occupancy for the ICU beds was 75% (273 occupied beds) and the average occupancy for the O2 bed was 50% (1134 occupied beds) (see table 1). The maximum occupancy occur on July 26, 2021, with 97% occupancy of ICU beds and 90% for O2 beds on July 7, 2021.

Similarly to the epidemiological curves, six waves were seen for O2 and ICU beds of various intensities (see figures 4a and 4b). Note that, for dedicated beds, the third and fourth waves were nearly identical. Moreover, until August, the number of dedicated beds continued to increase and did not change as much as the occupied bed curves. These two events highlight the government’s continued attempts to distribute beds.

**Figure 4:**
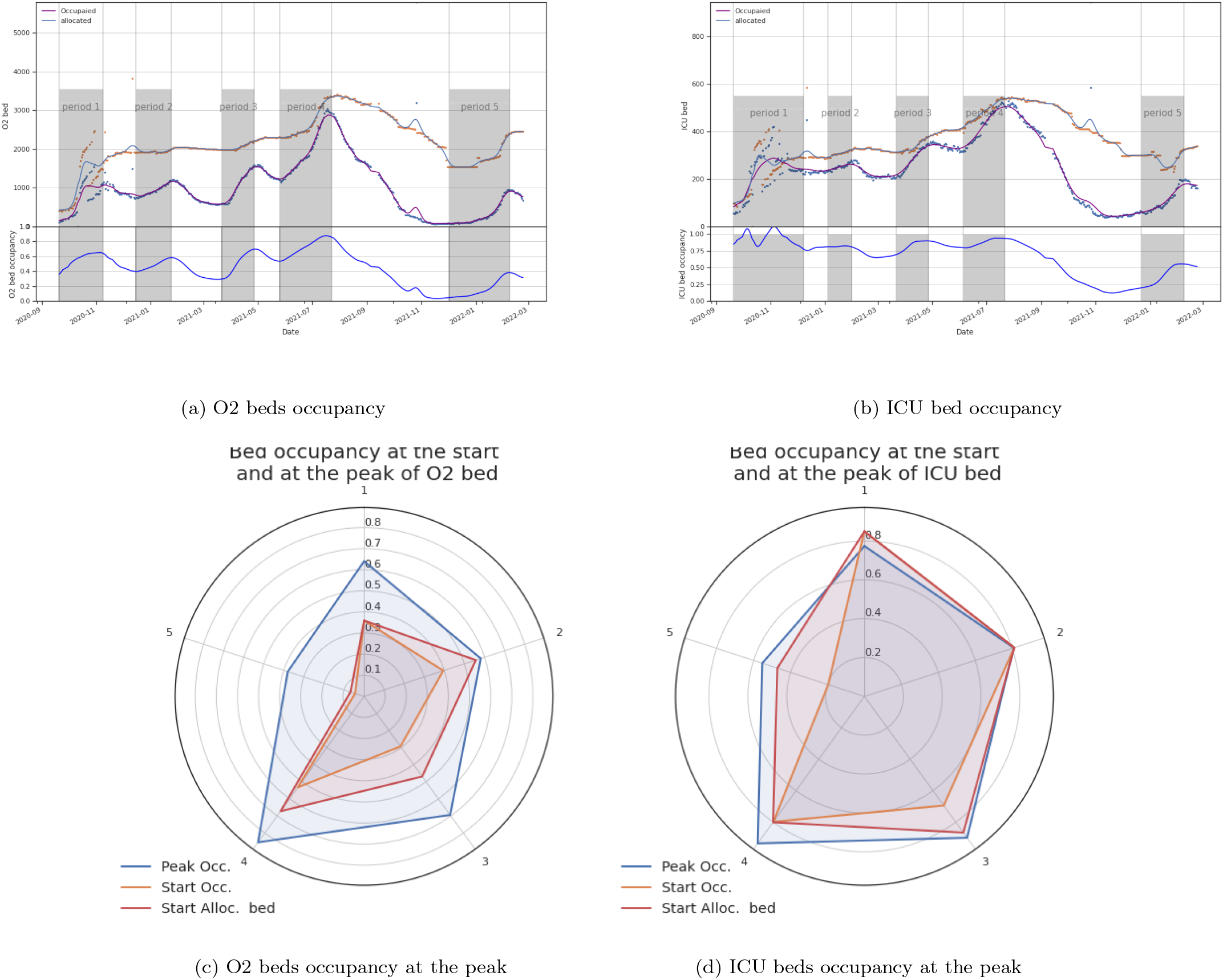
Bed occupancies for O2 beds (figures 4a and 4c) and ICU beds (figure 4b and 4d) from March 2020 to February 2022. We observe six waves for O2 and ICU beds of various intensities (see figures 4a and 4b). There is very small difference between occupancy at peak and at the beginning of the each wave for the ICU bed.

The occupancy of ICU beds shows little change and stays near to 80% except for the third and fourth waves, when it exceeds 90%, indicating that ICU services remained under pressure during the four first waves (see figures 4d), even though it has been able to adapt to the increasing severity of the epidemic.

In contrast to the ICU bed, the O2 bed occupancy curve oscillates and ranged around 50%, except for the third and notably the fourth waves, where it reached a maximum of 90% on July 7, 2020 (see figures 4c).

We observe that occupancy at the start of the occupied bed wave was less than at the start of the dedicated bed for O2 beds. It was, however, roughly equal for ICU beds (see figure 4d). This discrepancy may be explained by the fact that ICU bed distribution during waves is based on demand with a quick response.

This observation is also supported by an evaluation of the time lag between the start of the waves of occupied and dedicated beds. Indeed, we find that the average time delay for ICU beds is 11.4 days (standard deviation = 15.7), with a maximum of 31 days for the wave 5. The reason for the long time delay is that the rate of bed growth in wave 5 was slow(see figure 7d). The wave 3-time delay lasted 18 days.

The average time delay between occupied et allocated O2 beds was 22.75 days (standard deviation = 7.5), with a maximum of 33 days for wave 2.

We also plotted the occupancy at the start of each wave, and for O2 and ICU beds (see figures 4c and 4d). We found that the occupancy at the start of the allocated bed waves was higher than the occupancy at the start of the occupied bed waves, especially for O2 beds. This demonstrates a time delay in bed re-allocation, particularly for O2 beds (see figures 5 and 6a). However, the difference in occupancy at the start of the two ICU bed waves (allocated and occupied beds) is small, except for the third wave, where it is 70% *versus* 90%.

**Figure 5:**
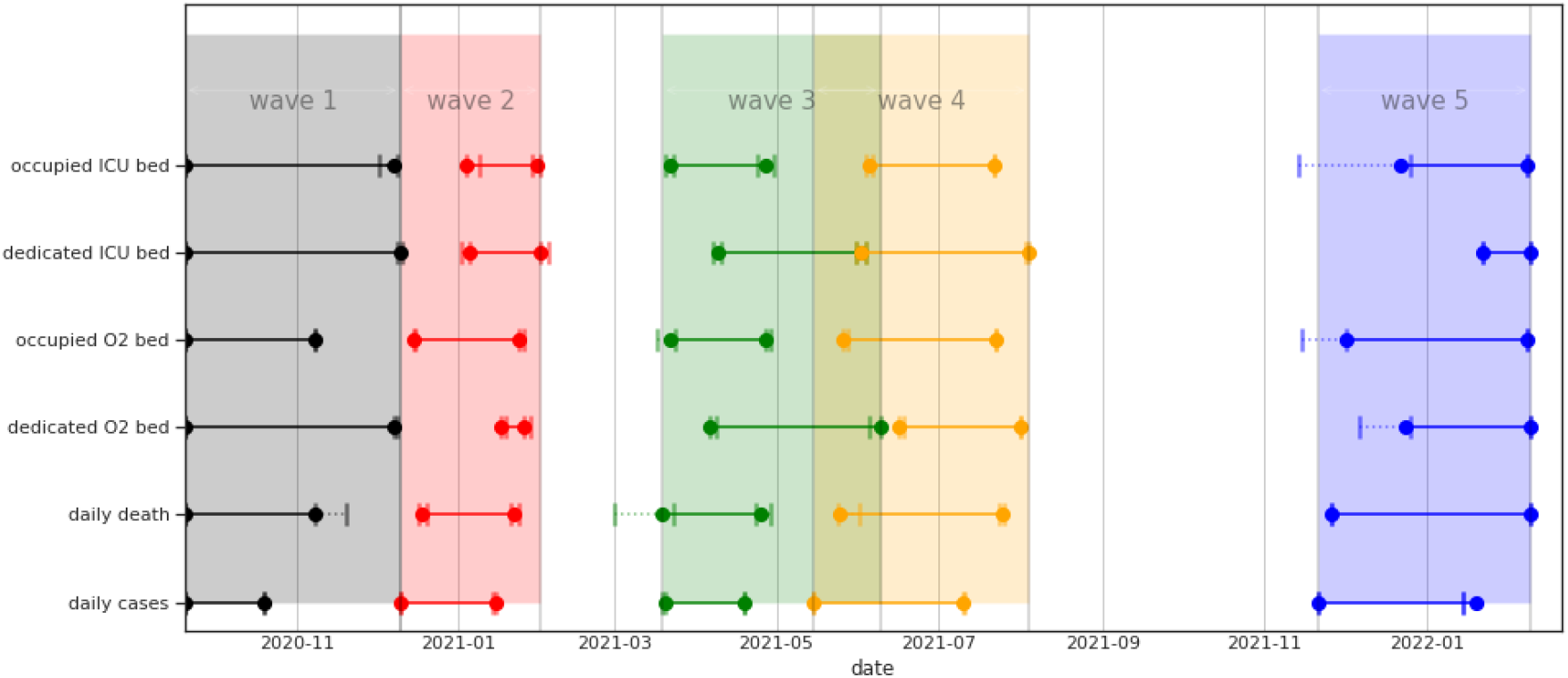
Time distribution of the different waves for daily cases, daily deaths and occupied and dedicated beds

**Figure 6:**
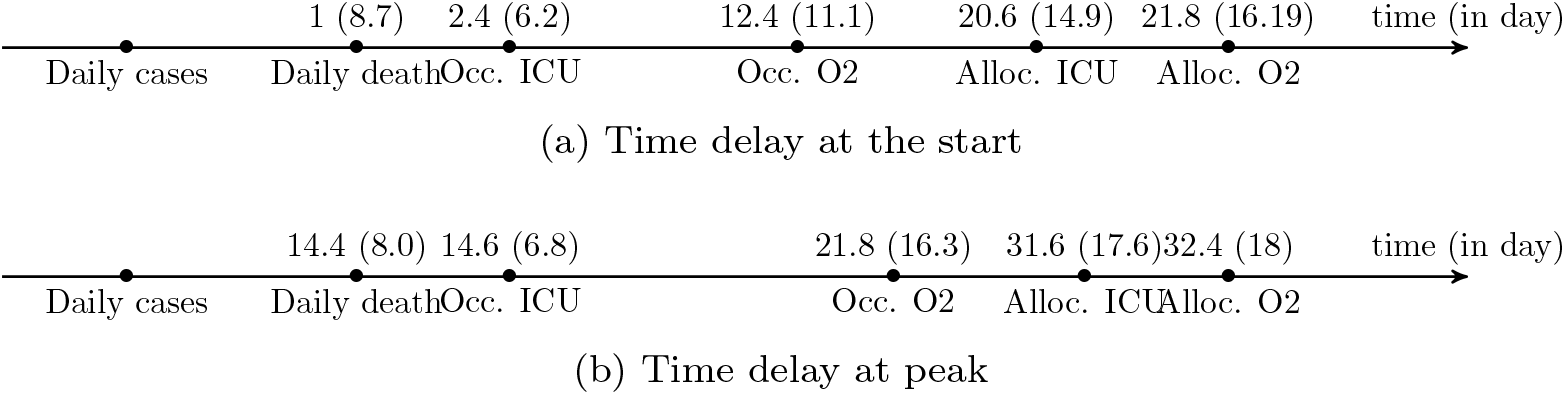
Mean delay and std at the start the peak by waves for for daily death, allocated and dedicated O2 and ICUbeds. The delay is calculated relatively to daily cases curves.

**Figure 7:**
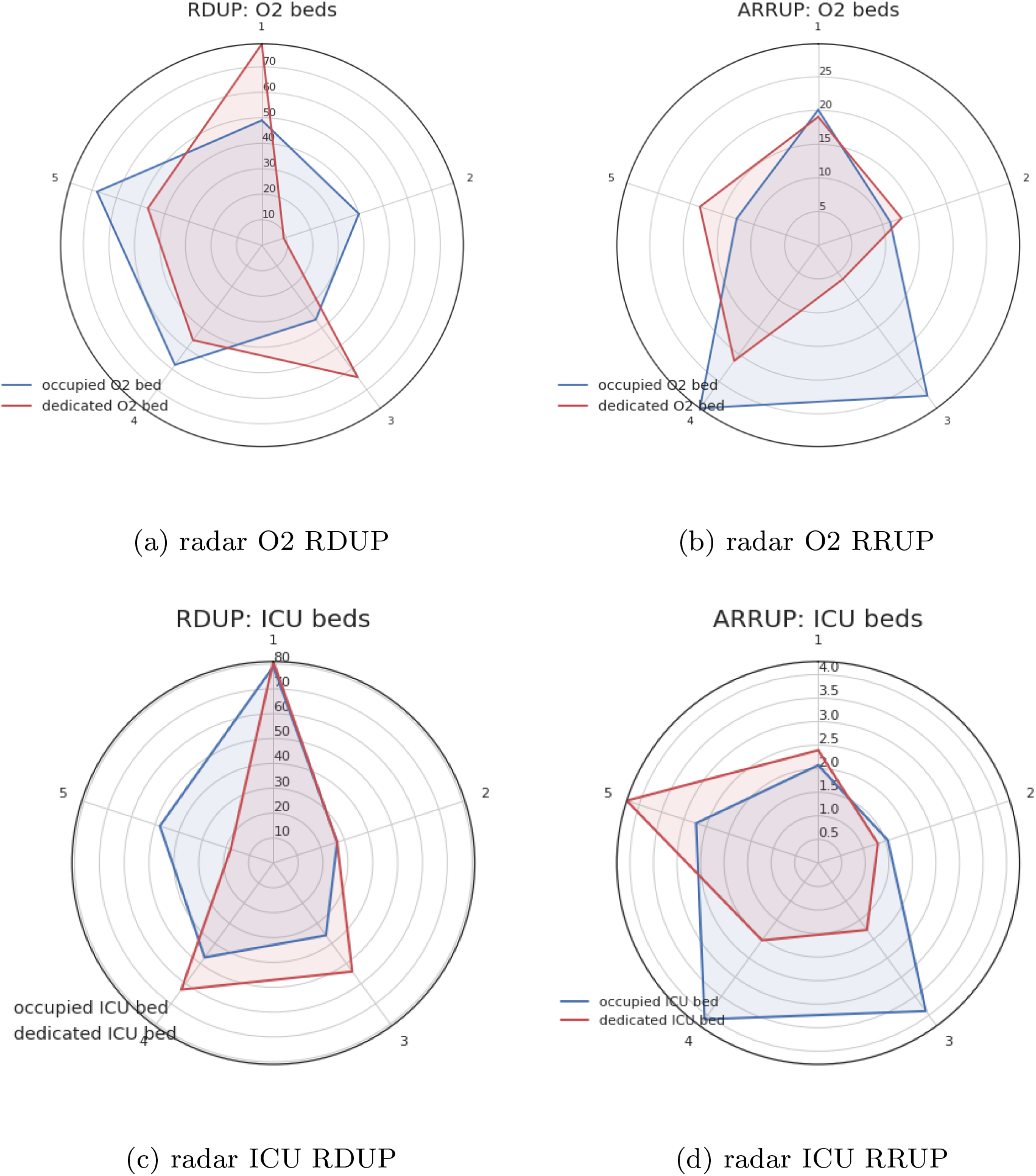
Ramp Duration Until the Peak (RDUP) and Ramp Rate Until the Peak (RRUP) for the five waves. On observe que les vague 3 et 4 ont ete les plus intense pour les lit occup’e (see figures 7b and 7d). Par contre les vague des lits allou’e ete plus longue mais moins intenses (see figures 7a and 7c).

**Figure 8:**
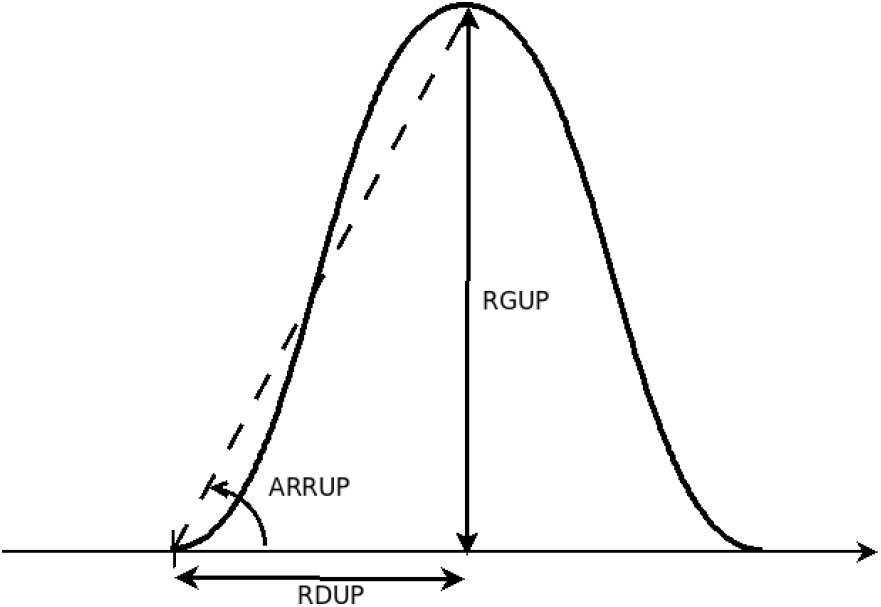
Flexibility index description: Ramp Duration Until the Peak (RDUP), Ramp Growth Until the Peak (RGUP) and the Ramp Rate Until the Peak (RRUP).

**Figure 9:**
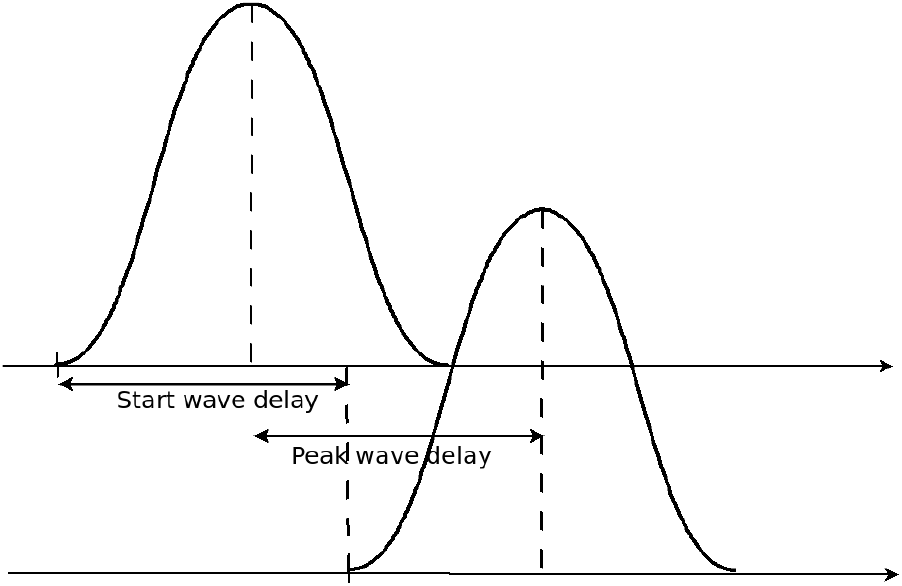
Time delay betwen two waves.

**Figure 10:**
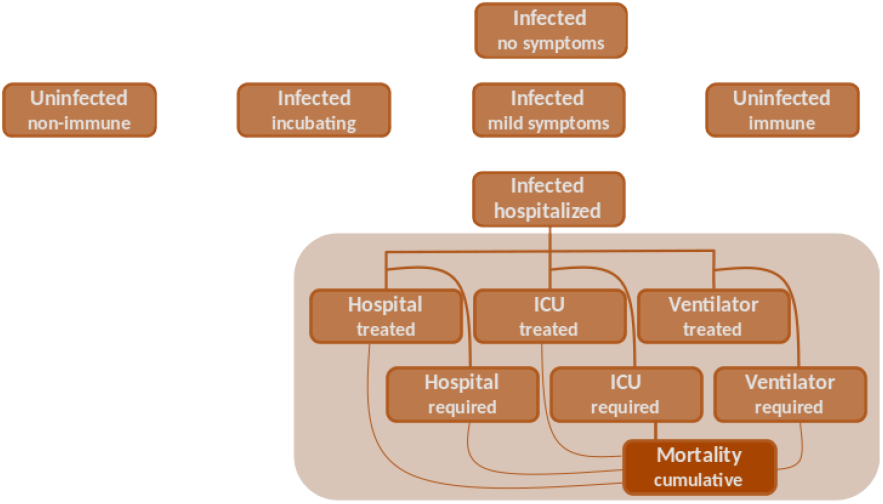
Coceptual view of the CoMo Model.

We then analysed the difference in the peak times (see figures 5 and 6b. The order of appearance of the peak waves appears to be as follows: daily case, daily death (with an average time delay of 14.4 days, std=8), occupied ICU beds (with an average time delay of 14.6 days, std=6.8), occupied O2 (with an average time delay of 121.8 days, std=16.3), dedicated ICU bed (with an average time delay of 31.6 days, std=17) It should be highlighted that the difference between daily death and ICU occupied, as well as dedicated beds, is extremely small (see figure 6b).

Furthermore, it appears that beds are still being allocated after the peak of occupancy, which lasts an average of 10.6 days (std=15) for ICU beds and 17 days (std=18.6) for O2 beds. The maximum time delay for ICU was observed for wave 3 where the time delay was 36 days at the pick, For O2 bed, we found that during wave 3, O2 beds continue to be allocated 43 days after the peak of the occupied bed’s wave (see figure 6b).

For all waves, we observe that the peak of deaths occurs before the peak of ICU bed occupancy. This mismatch might be caused by patient bed occupancy periods or by a carry-over of deaths from the previous wave to the beginning of the new wave. However, the mean time spent in ICU is 7.33 days (ssd=5.88) for dead and 6.83 days (ssd=5.45) for recovery and therefore, cannot explain this discrepancy. An alternative could be the large values of standard deviation, 5.88 days for death and 5.45 days for recovery. Another explanation could be the high number of death occurring outside the hospital.

To investigate the duration and strength of each wave, we studied the evolution of the Ramp Duration Until the Peak (RDUP) and the Ramp Rate Until the Peak (RRUP).

Except for waves 1 and 3 for O2 beds, the RDUP for beds occupied was longer than the allocated beds (see figure 7). Furthermore, except for wave 5, we found the RRUP was higher for occupied beds.

In the example of the occupied O2 bed, we can see that it rapidly increases during wave 4 (RDUP=58 days and a massive RRUP=29.8). Furthermore, wave 3 exhibits rapid growth (RRUP= 27.5) during a short length of time (RDUP=36 days). The dedicated bed, on the other hand, has developed slowly and gradually.

For ICU beds, occupied has had rapid growth in a short time. Dedicated beds, on the other hand, display slower growth over a longer period (except for wave 5). The higher increase was done for wave 3 and for where RRUP= 4.1 (RDUP=47 days) for wave 4 and 3.8 for wave 3(RDUP=36 days).

In a short time, the number of ICU beds occupied has increased rapidly. Dedicated beds, on the other hand, exhibit slower growth over a longer length of time (except for wave 5). The larger increase was performed for wave 3 where RRUP= 4.1 (RDUP=47 days) and for wave 4 where RRUP= 3.8 (RDUP=36 days).

## 4 Discussion

The analysis of COVID-19 healthcare data has revealed an opportunity to better understand healthcare performance in stressful circumstances. We were able to assess the overall responsiveness of the Tunisian public hospital system by assessing flexibility and bed margin. Decision makers will be able to assess their response capabilities in the case of a current pandemic thanks to this research.

Decision-makers in public health should respond, especially in emergencies where the system is forced to manage critical and urgent risk ((Didier, 2016)). This is accomplished through the proactive deployment of NPI measures such as curfews, lockdowns, masking requirements, or school and administrative closures to avoid or lessen dangers.

These decisions are frequently influenced by financial restrictions. For example, during the first wave in Tunisia between March and June 2020, rigorous confinement for three weeks was implemented. As a result of these steps, the number of instances has decreased. However, these measures resulted in a drop of at least 4.4 percent of GDP and a 21.6 percent unemployment rate, according to UNDP ((2020)).

Economic constraints force developing countries, such as Tunisia, to choose between two approaches to crisis management: an economically costly strategy targeted at minimizing deaths or a policy aimed at avoiding excess deaths owing to health-system saturation.

Tunisia, beginning in July 2020, seems to be characterized by incentives aimed at avoiding hospital overcrowding. Apart from school closure decisions, we found in our study that required NPI decisions were poorly implemented and frequently time delayed. During the same period, we noticed that people were aware of the severity of the epidemic by taking precautionary measures such as wearing masks. This was most clear during the second wave, between September and October 2020, when the decision to implement a nationwide lockdown was made after the epidemiological peak. During this same period, we observed a significant increase in the use of masks among individuals.

During a health crisis, the decision-maker must determine when, how much, and where to commit the necessary resources to manage the epidemic-resources are either human (human resources reallocated from one service to another) or material (bed allocations to COVID-19). The choice is additionally hampered by the lack of knowledge on (i) the current and future state of the epidemic, and (ii) the health system’s reactivity in terms of the time delay in executing the remedies decided upon Tabuteau ((2008)).

We found a negative time delay between occupancy and assigned beds at the start of the waves, as well as a positive time delay at the peak. This implies that the authorities kept allocating beds long after the peak of the occupied bed wave had passed. These time delays were most obvious for O2 beds and during peak times. This sustained effort contributed to reducing pressure during the second and, especially, fourth waves, when the rate of increase in O2 beds and ICU was substantially slower than the rate of increase in daily patients. Furthermore, the overlap in allocated beds between the third and fourth waves may be explained by the fact that these two epidemiological waves were close together, with the third wave’s peak of daily cases on April 19 and the start of the fourth wave on May 15.

Moreover, we observe that the higher occupancy of ICU beds at the beginning of each wave has likely resulted in a quicker rate of ICU bed allocation (mean 4.0 days, std=9.48) compared to O2 beds O2 beds (mean 17.25 days, std=13.72). This discrepancy in time delay might be explained by the fact that decisions were made primarily based on bed occupancy rather than a wave start evaluation.

Such time delays could be the result of a precautionary principle based on a lack of information or trust in data on the evolution of the epidemic.

## 5 Conclusion

By comparing the NPI activities performed and the response in terms of daily detection cases, we were able to recreate the history of the evolution of the epidemic in Tunisia. We discovered that, except for the first wave, the NPI activities were poorly implemented, despite the importance of the decisions made. This failure to follow the constraints, along with the presence of increasingly contagious variations, allowed for the creation of larger waves.

The analysis of bed occupancy revealed that there was a delay in the allocation of new beds at the start of the pandemic, as well as in the reallocation of beds at the peak. At its peak, this delay was more than 30 days on average for O2 and ICU allotted beds. These delays demonstrate the health system’s slow response to the epidemic’s progression at the start of the waves and prudence at the peak.

As a perspective, the information presented here could be useful in comparing how effective different health care systems are at responding to pandemics in terms of response time, particularly between developed and developing countries.

## Data Availability

The database can be found on GitHub:
https://github.com/slimane66/bedOccupancyTunisia.git

## A Correlation analysis

## B Used database

**Figure 11:**
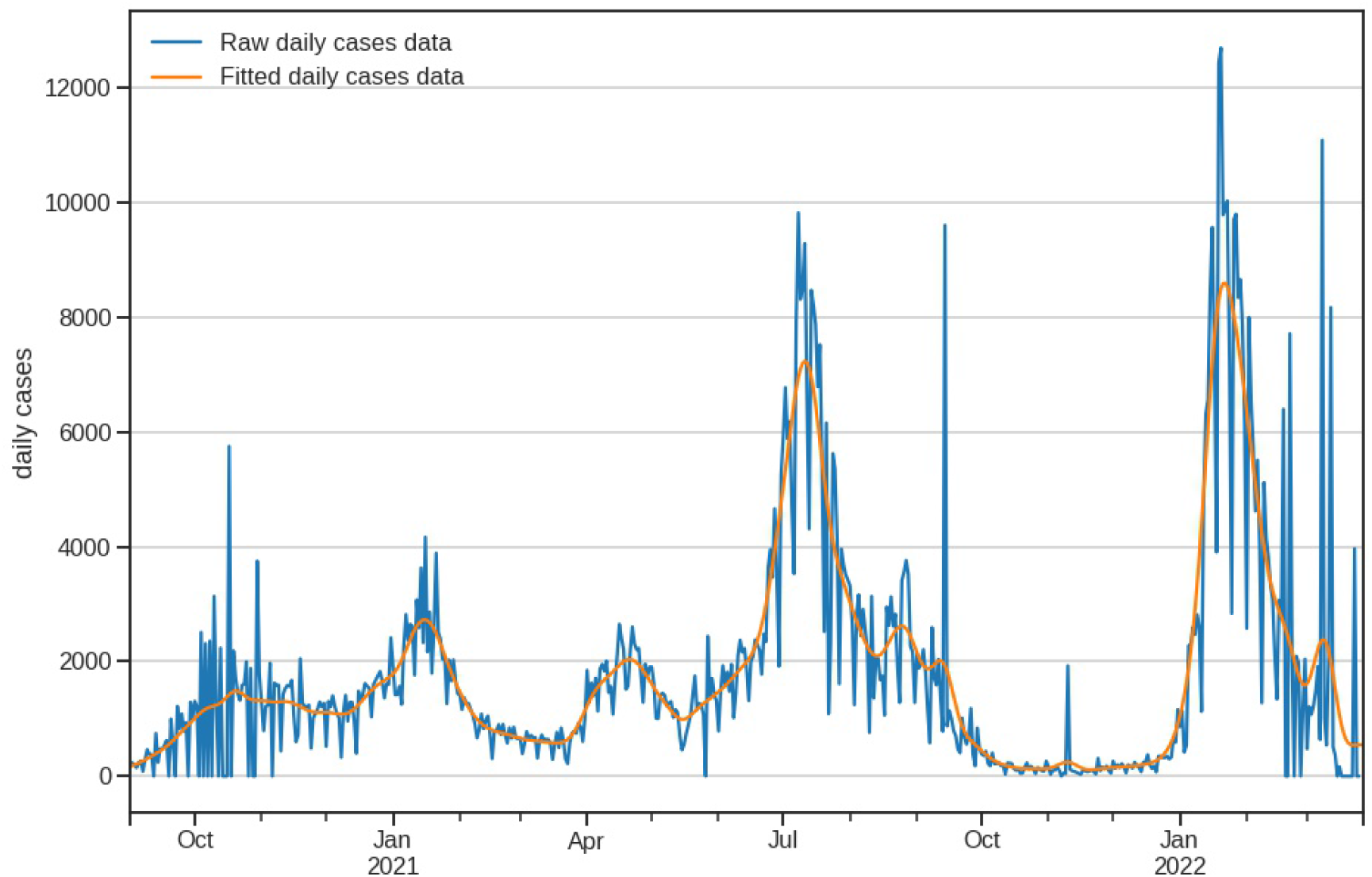
Daily cases: data and fitted curve. Data was fitted using a linear convolution one dimension kernel, *Box*1*DKernel*(*N*) with *N* = 7 days. (c) Occupaied O2 beds (d) Occupaied ICU beds

**Figure 12:**
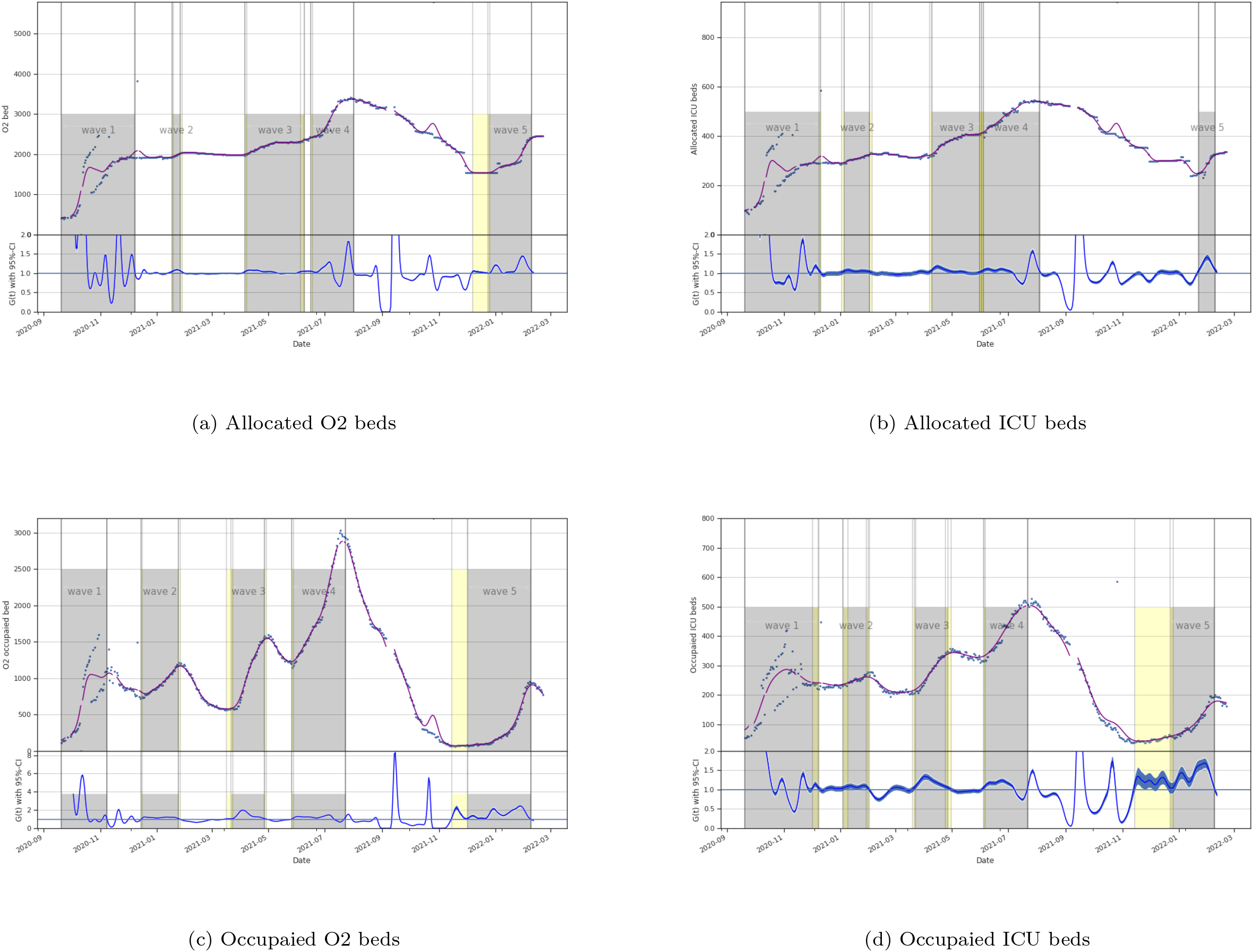
Beds evolution by time with their corresponding *G*_*t*_.

**Figure 13:**
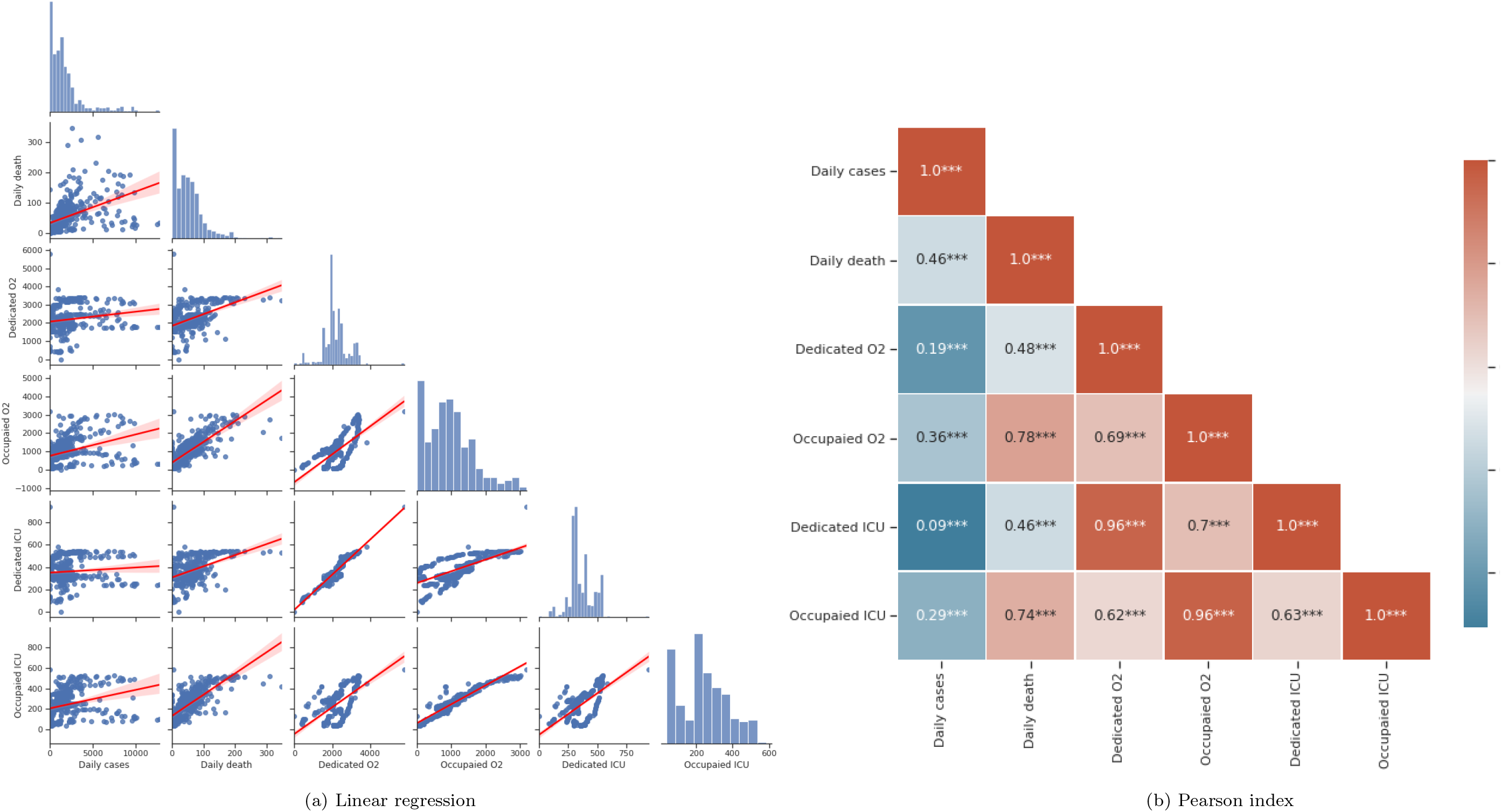
Pairplot and Pearson index (*** stand for *p*-value *<* 0.001) for bed occupancy and epidemiological curve. It appears that occupied and dedicated beds are strongly correlated (*R*2 *>* 0.8, *p - value <* 0.001). Moreover, occupied beds are strongly correlated to daily cases and daily deaths. However, dedicated bed a less correlated to daily cases and daily deaths (*R*2 *<* 0.6, *p - value <* 0.001).

**Figure 14:**
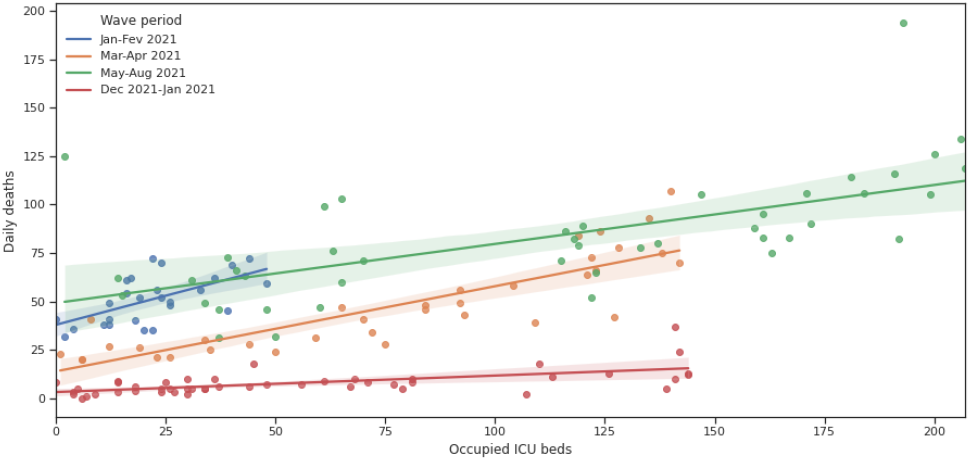
Linear regression between ICU bed occupancies and daily death for wave 2,3,4 and 5. The intercept is respectivley 0.56^*\*\**^0.42^*\*\**^, 0.33^*\*\**^, 0.08^*\*\**^ (*R*2 = 0.34, 0.74, 0.465, 0.387)

## Footnotes

1 https://dataforgood.facebook.com/

2 https://www.healthdata.org/covid/

3 http://www.ins.tn/

4 https://covid19.who.int/

5 https://nextstrain.org/ncov/

## References

R. Aguas, L. White, N. Hupert, R. Shretta, W. Pan-Ngum, O. Celhay, A. Moldokmatova, F. Arifi, A. Mirzazadeh, H. Sharifi, K. Adib, M. N. Sahak, C. Franco, and R. Coutinho. Modelling the COVID-19 pandemic in context: an international participatory approach. BMJ Global Health, 5(12):e003126, dec 2020. ISSN 2059-7908. doi: 10.1136/bmjgh-2020-003126. URL https://gh.bmj.com/lookup/doi/10.1136/bmjgh-2020-003126.

R. Bekker, G. Koole, and D. Roubos. Flexible bed allocations for hospital wards. Health Care Management Science, 20(4):453–466, 2017. ISSN 13869620. doi: 10.1007/s10729-016-9364-4.

A. Cori, N. M. Ferguson, C. Fraser, and S. Cauchemez. A new framework and software to estimate time-varying reproduction numbers during epidemics. American Journal of Epidemiology, 178(9):1505–1512, 2013. ISSN 00029262. doi: 10.1093/aje/kwt133.

H. Didier. Maintenant ou trop tard, sur le phenomene de l’urgence. Denoel, 2016. ISBN 2207252256.

O. Diekmann, J. A. Heesterbeek, and J. A. Metz. On the definition and the computation of the basic reproduction ratio R0 in models for infectious diseases in heterogeneous populations. Journal of Mathematical Biology, 28(4):365–382, 1990. ISSN 14321416. doi: 10.1007/BF00178324.

L. V. Green and L. G. V. How Many Hospital Beds? Inquiry : a journal of medical care organization, provision and financing, 1(4):763, 2002. ISSN 0046-9580 (Print). doi: 10.1136/bmj.1.5021.763-a.

J. Hadfield, C. Megill, S. M. Bell, J. Huddleston, B. Potter, C. Callender, P. Sagulenko, T. Bedford, and R. A. Neher. Nextstrain: realtime tracking of pathogen evolution. Bioinformatics, 34(23):4121–4123, dec 2018. ISSN 1367-4803. doi: 10.1093/bioinformatics/bty407. URL https://doi.org/10.1093/bioinformatics/bty407.

T. Hale, S. Webster, A. Petherick, T. Phillips, B. K. last Updated, and U. 2020. Oxford COVID-19 government response tracker (OxCGRT), 2020. URL https://covidtracker.bsg.ox.ac.uk/.

T. L. Lash, T. J. VanderWeele, S. Haneuse, and K. J. Rothman. Modern epidemiology. Wolters Kluwer, 4 edition, 2020. ISBN 9781451193282.

O. Ouchetto, A. Drissi Bourhanbour, and M. Boumhamdi. Effectiveness of containment measures to control the spread of COVID-19 in North Africa. Disaster Medicine and Public Health Preparedness, pages 1—-5, 2020. ISSN 1938744X. doi: 10.1017/dmp.2020.314.

T. Robitaille, Astropy Consortium, and M. Urbina-Fuentes. Astropy: A community Python package for astronomy, 2013. ISSN 25902296. URL https://docs.astropy.org/en/stable/https://doi.org/10.1051/0004-6361/201322068.

C. Sohrabi, Z. Alsafi, N. O’Neill, M. Khan, A. Kerwan, A. Al-Jabir, C. Iosifidis, and R. Agha. World Health Organization declares global emergency: A review of the 2019 novel coronavirus (COVID-19). International Journal of Surgery, 76:71–76, 2020. ISSN 1743-9191. doi: https://doi.org/10.1016/j.ijsu.2020.02.034. URL https://www.sciencedirect.com/science/article/pii/S1743919120301977.

D. Tabuteau. La decision en sante. Sante Publique, 20(4):297–312, 2008. ISSN 09953914. doi: 10.3917/spub.084.0297.

UNDP. Impact Economique du COVID 19 en Tunisie. Technical report, Republique Tunisienne, PNUD, 2020. URL http://www.mdici.gov.tn/impact-economique-de-la-pandemie-du-covid-19-sur-leconomie-tunisienne-

G. Van Rossum and F. L. Drake. Python 3 Reference Manual. CreateSpace, Scotts Valley, CA, 2009. ISBN 1441412697.

WHO. WHO announces COVID-19 outbreak a pandemic 2020, 2020. URL

